# Nano short peptide nutrition intervention on the prognosis of patients with COVID-19

**DOI:** 10.1101/2020.06.03.20083980

**Authors:** Zelin Zhang, Lin Yang, Tianping Zhang

## Abstract

**Background:** The continuous outbreak of COVID-19 poses a devastating threat to the global public health, and there’s no special therapeutic drugs. This paper is to explore the effect of high fiber whey short peptide enteral nutrition on the prognosis of patients with COVID-19, to find ways to prevent patients from progressing to severe illness, and to reduce the harm of epidemic situation to human beings.

**Methods:** The course of fighting against COVID-19 in our hospital for 49 days was reviewed. Three nutritional interventions including five-step nutrition treatment, ^46^early intervention of whole protein, and high fiber whey short peptide intervention were conducted consecutively.The effect of high fiber whey short peptide on nutrition, immune function and prognosis of patients with COVID-19 was compared with that of whole protein intervention.

**Results:** High fiber whey short peptide group when compared with whey protein group, can significantly improve prealbumin in patients with covid-19, improve the negative nitrogen balance of patients, and reduce the average period of turning negative by 39.06%.

**Conclusion:** High fiber whey short peptide can significantly improve the recovery speed of patients with covid-19.

**Funding:** none

## Introduction

The absolute number of T lymphocytes decreased in almost all patients with COVID-19, and the immune function of patients with covid-19 is generally low.^21-33^ The prognosis of patients with COVID-19 largely depends on their own immune regulation. If the speed of acquired immunity is lower than the speed of virus development, the condition will deteriorate rapidly. ^23^ Improving the immunity of patients with COVID-19 and killing sars-cov-2 by the human body’s own immune function have become the key factors of current anti epidemic.In this paper, 208 patients with COVID-19 who were diagnosed in our hospital from January 30, 2020 to March 17, 2020 and received early nutrition intervention were reviewed. The effects of early whole protein intervention and high fiber whey short peptide intervention on nutrition index, immune function and prognosis were studied.

## Data and methods

### Clinical data

All patients were recruited from Yichang Central People’s Hospital in Hubei province.Since January 29, 2020, all inpatients with COVID-19 have been given five-step treatment of enteral nutrition intervention.^46^ Since February 8, 2020, the regimen shifted to whole protein prophylactic enteral nutrition. Methods: On the basis of conventional diet, all common type patients, took orally 50g whole protein (Homogenate diet, including milk protein and soybean protein)as enteral nutrition every day. According to the condition of severe and critical patients, individualized nutrition programs were developed in a total of 206 patients. Since February 22, 2020, 49 patients with slow recovery after preventive nutrition intervention with whole protein,received enteral nutrition intervention with high fiber whey short peptide instead of whole protein(Brand: fanting,batch No.: 20191005, 20200223,specification: 200ml / bottle, nutrition ingredients in each bottle: 8g nano short peptide, 1g amino acid, 3.0g nano soybean dietary fiber, 4.0g oligofructose).Methods: On the basis of traditional diet,daily oral administration of high fiber whey short peptide was given as enteral nutrition, with no more whole protein. Among them, 20 patients each drank 3 bottles a day, 7 patients each drank 2 bottles a day,and 22 patients each drank 1 bottles a day.Those taking 2-3 bottles per person per day were recruited as the study subjects. 12 patients were excluded from the study for the duration less than 2 days or their light or severe types. 15 common patients (2 of 2 bottles, 13 of 3 bottles) were used as the treatment group, to study nutritional status and immune indicators. Among 206 confirmed patients with covid-19, those of light, severe, and critical types, or death, or those with non continuous intervention till discharge or with incomplete detection indexes were excluded. According to the sex and age of the treatment group, 51 common cases were matched as the control group. There was no significant difference between general situation of the two groups(p>0.05)(table1)

**Table 1:** Comparison of general data between two groups of patients

| Index | Treatment group(n=13) | Control group(51) |
| --- | --- | --- |
| Gender(man/woman) | 7/8 | 24/27 |
| Average age/year | 47.8±17.10 | 520.71±14.51 |
| Age minimum/maximum | 29/83 | 18/68 |
| COVID-19typing | Ordinary type | Ordinary type |
| Underlying diseases/cases | 33.33% | 29.41% |
| Respiratory system | 1 | 1 |
| Cardiovascular system | 2 | 7 |
| Cerebral vascular disease | 1 | 1 |
| chronic hepatitis B | 0 | 5 |
| hyperuricemia | 0 | 1 |

## Methods

As for data of treatment group and control group,according to the duration of the disease,the acquisition standard took 3-7 days as the node.^16.17^ Through the contrast between groups and before and after intervention, the therapeutic effect of high fiber whey short peptide on patients with COVID-19 was studied.

The data was input into spss19.0 statistical software for processing, and each variable is represented by 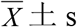. T test is used to compare the mean of measurement variables. The counting data is checked by X^2^. P < 0.05 has statistical significance.

## Results

### Clinical nutrition analysis of patients with COVID-19

In this study, 206 patients with COVID-19 were treated with whole protein. These patients include mild, common, severe, critical and dead patients. At the time of admission, the proportion of people whose hemoglobin, total protein and albumin were lower than normal was relatively high. 151 of the 152 people who tested prealbumin were lower than normal. The average value of 151 human prealbumin was 149.08mg/1. 99.34% of the patients were in a negative nitrogen balance state. The mean value of prealbumin was 147.36mg/lin 140 patients with no obvious abnormal liver function, and was 171.05mg/1 in 11 patients with obvious liver function abnormality. The screening criteria of liver function abnormality:

AST >100u/L,ALT >100u/L,STB > 42.8 μ mol/L,GGT >100u/L, ALP >275u/L, the decrease of prealbumin in these patients had no significant relationship with liver function damage(Table 2).

**Table 2.**
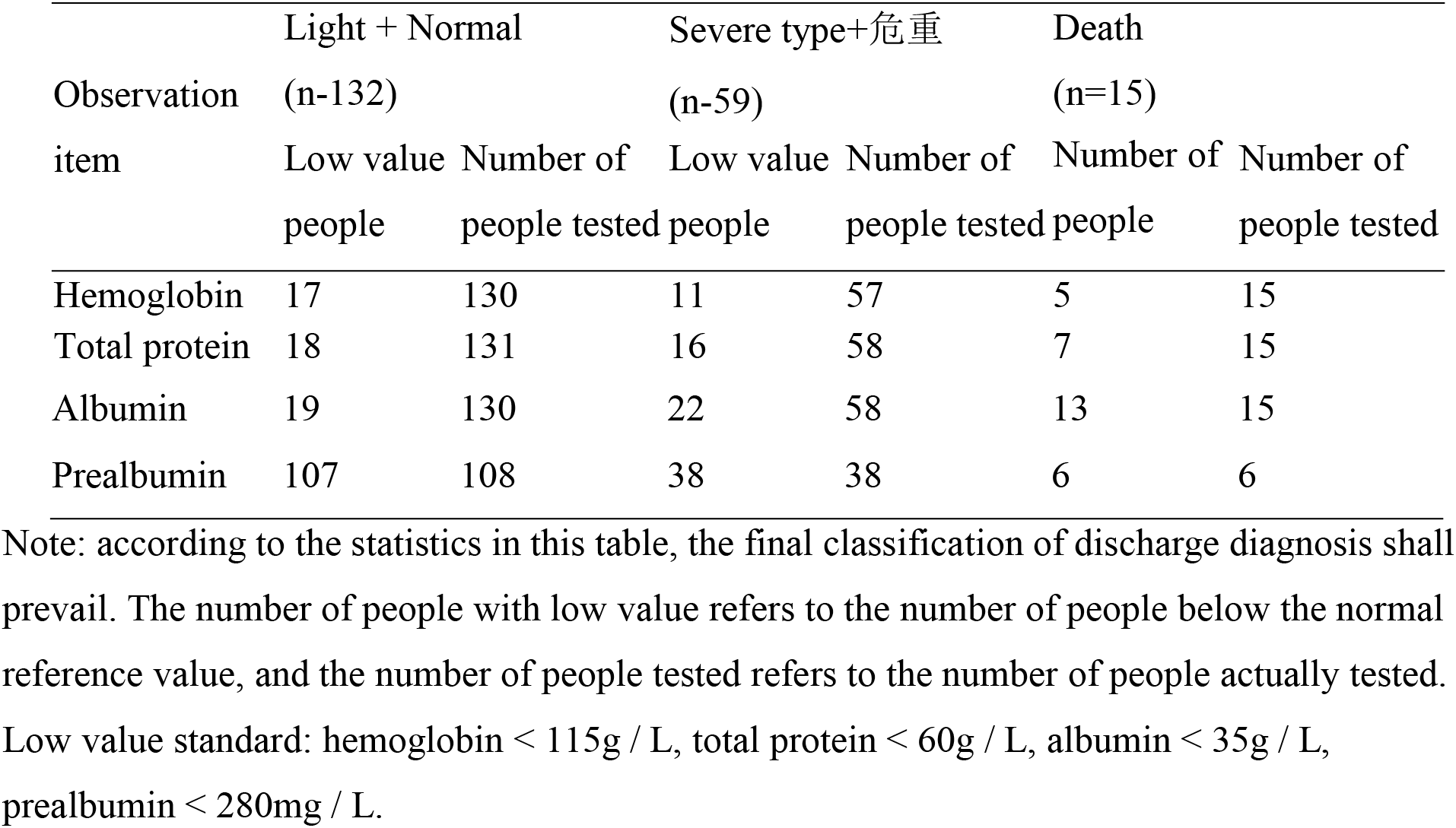
Nutritional status on admission.

### Comparison of prealbumin between two groups

Throughout the course of the disease, prealbumin in both groups was lower than normal. In the control group, prealbumin didn’t differ between adjacent nodes(P>0.05), and was significantly higher at the discharge node than at the admission node(p=0.000).In the treatment group, prealbumin descended at the first node(p=0.721), and was significantly higher at the second node than at the first node (P = 0.004),and at the discharge node than at the admission node(p=0.001).The four nodes of the treatment group and the control group were compared, at the second and discharge nodes, prealbumin at the treatment group was no higher than that in the control group. There was no significant difference between the two groups (P > 0.05).(Table 4)

**Table 4.** Comparison of prealbumin between two groups mg/L

| Node | Treatment group(n=15) | Control group(n=51) | p value |
| --- | --- | --- | --- |
| Be hospitalized | 170.93±56.12 | 141.5±46.98 | 0.065 |
| First node | 161.50±89.10 | 180.33±73.64 | 0.541 |
| Second node | 255.66±51.17 | 211.55±75.54 | 0.141 |
| Leave hospital | 265.98±47.35 | 241.25±76.67 | 0.325 |

Note: P value is the p value of the comparison between two groups of corresponding nodes.

**Table 5.** Ratio of prealbumin lower than normal in two groups

| Node | Treatment group(n=15) | Control group(n=51) |
| --- | --- | --- |
| Be hospitalized | 11/11 (100%) | 42/42 (100%) |
| First node | 8/8 (100%) | 25/26 (96.15%) |
| Second node | 5/8(62.5%) | 18/20 (90.00%) |
| Leave hospital | 5/11(45.45%) | 20/30 (66.67%) |

**Figure 1:**
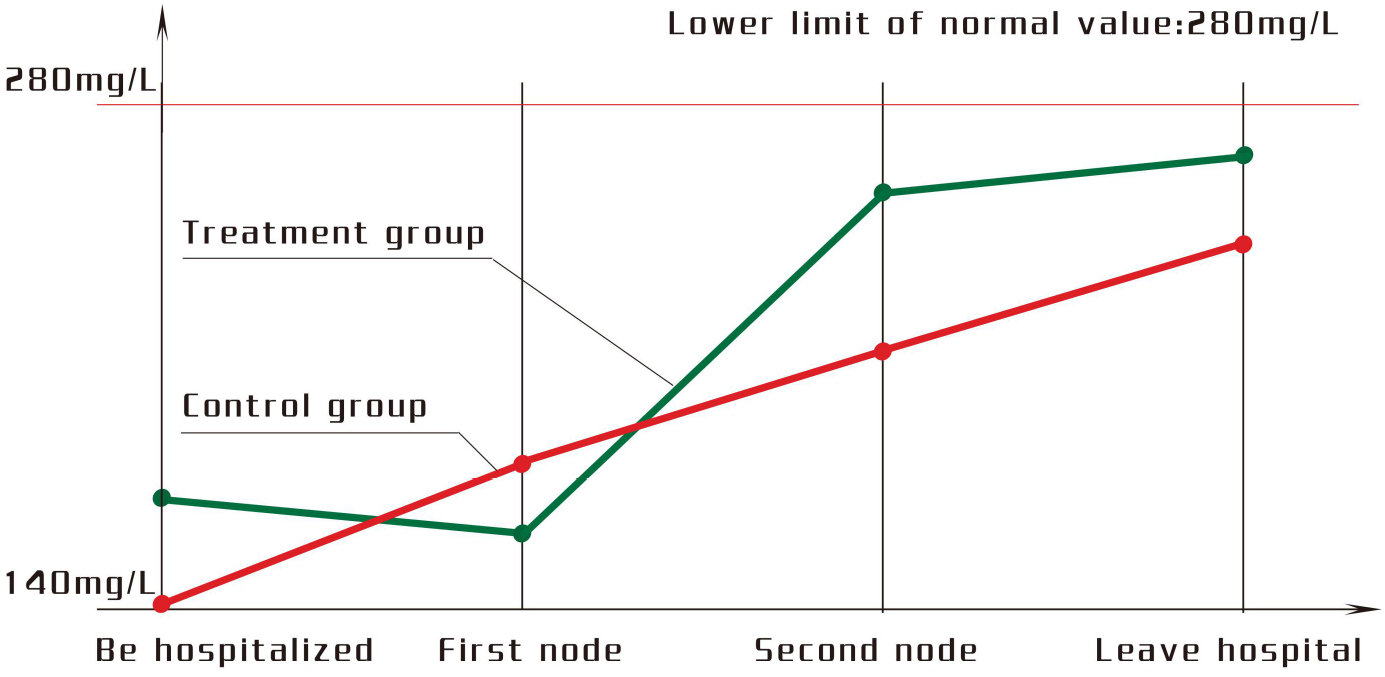
Numerical changes curve of prealbumin in two groups of patients

**Figure 2:**
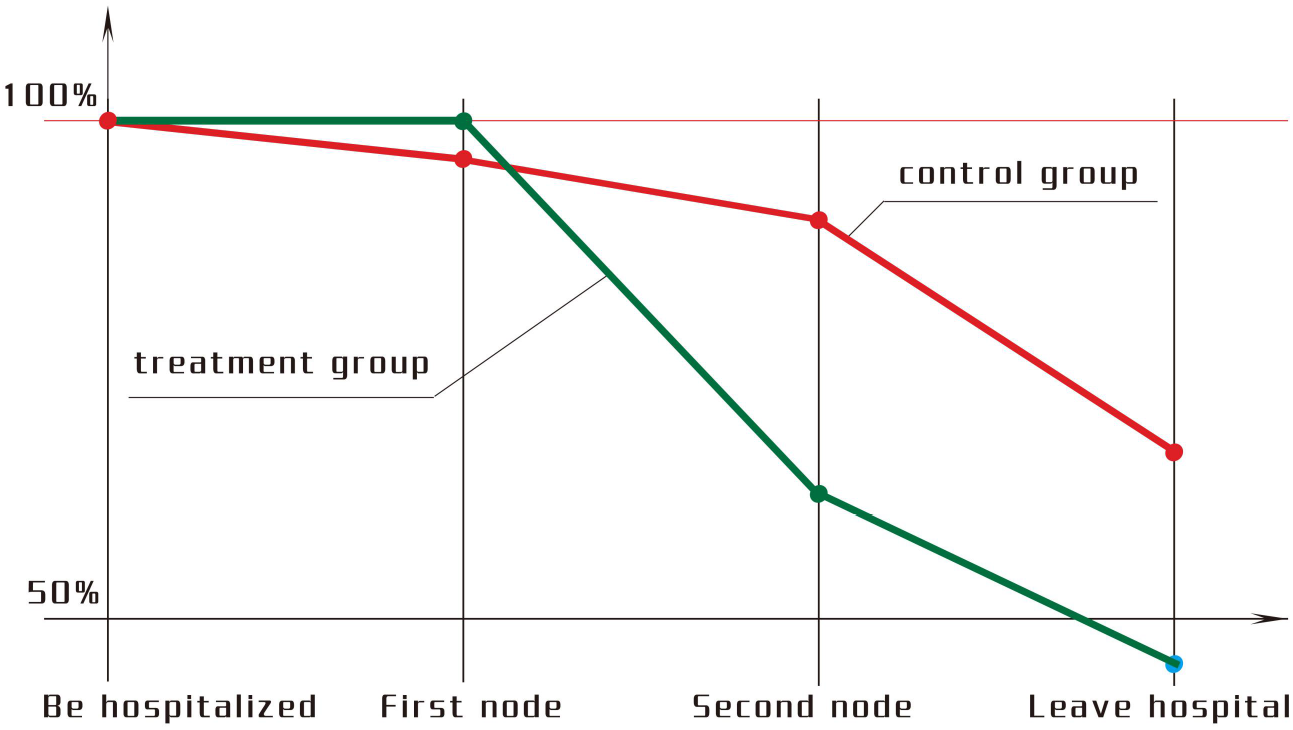
Percentage change curve of patients prealbumin lower than normal in two groups

### Comparison of lymphocyte between two groups

Throughout the course of the disease,the number of lymphocytes in the treatment group and the control group increased gradually. There were significant differences between the two groups from admission node to the second node and to discharge node (Treatment groupP = 0.001/0,Control group P = 0.018/0).The number of lymphocytes in the treatment group was higher than that in the control group. There was no significant difference between the two groups (P > 0.05)(Table 6).

**Table 6.**
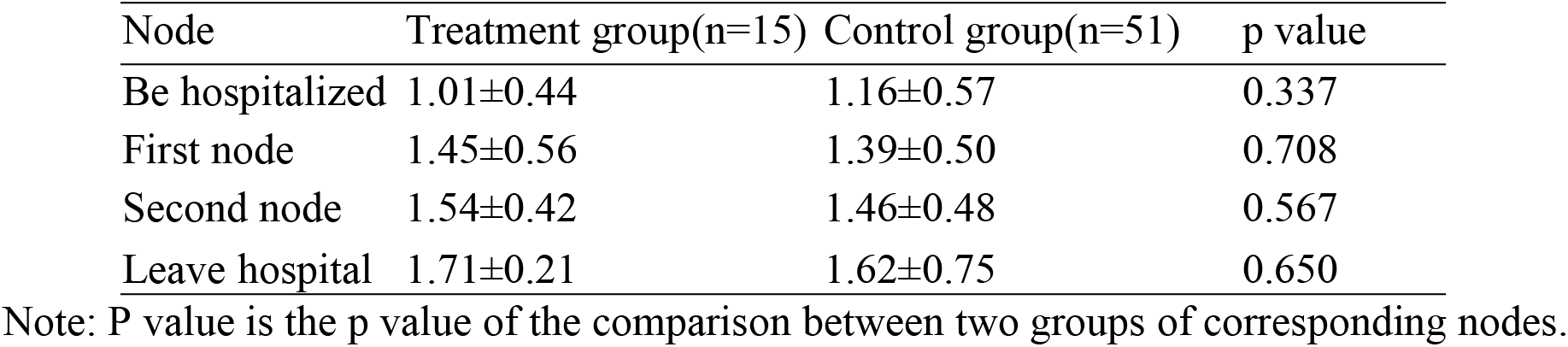
Comparison of lymphocyte number between two groups of patients × 10^9^/L

| Node | Treatment group(n=15) | Control group(n=51) | p value |
| --- | --- | --- | --- |
| Be hospitalized | 1.01±0.44 | 1.16±0.57 | 0.337 |
| First node | 1.45±0.56 | 1.39±0.50 | 0.708 |
| Second node | 1.54±0.42 | 1.46±0.48 | 0.567 |
| Leave hospital | 1.71±0.21 | 1.62±0.75 | 0.650 |

Note: P value is the p value of the comparison between two groups of corresponding nodes.

**Table 7.**
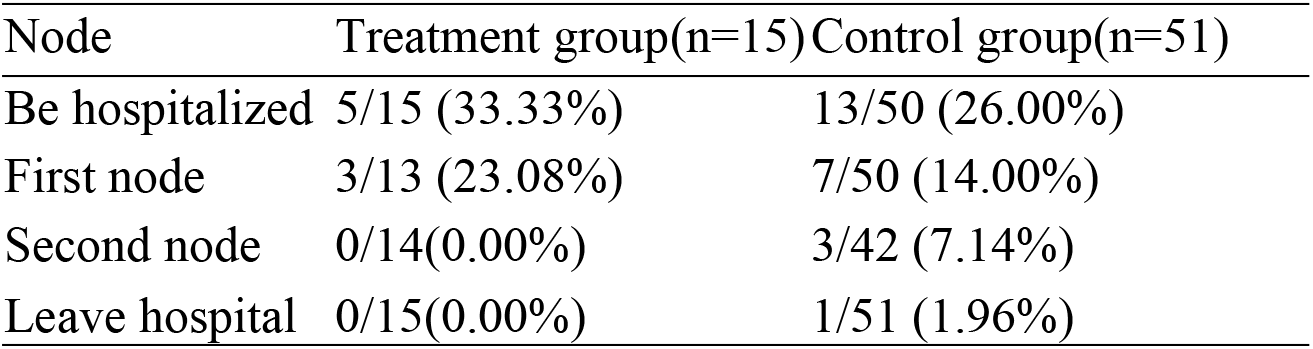
Ratio of lymphocyte below normal value in two groups × 100%

In the control group, the percentage of lymphocytes and the percentage of lymphocytes below normal were stable. The curve of lymphocyte number in treatment group was similar to that in control group, and the percentage change curve of the number of lymphocyte below normal value in the treatment group decreased significantly at the second node.

**Figure 3:**
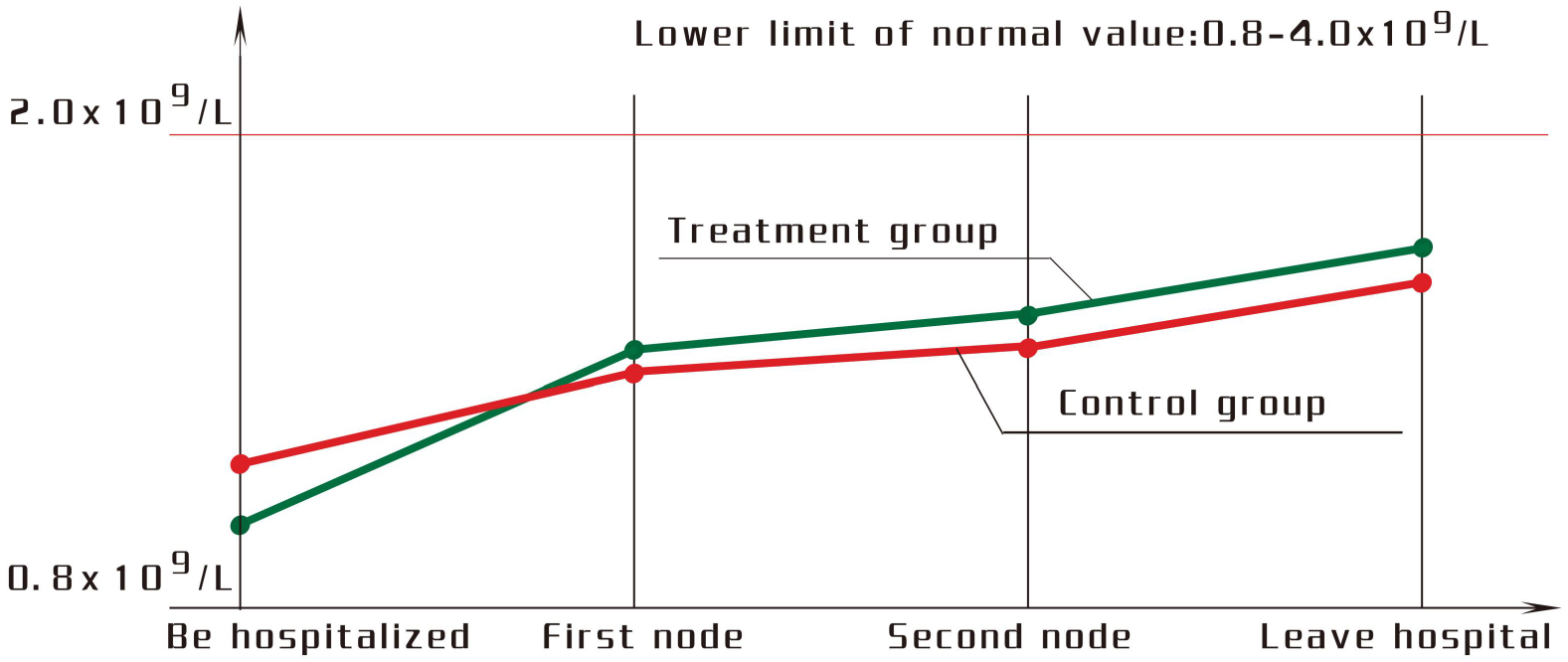
the change curve of the number of lymphocytes in two groups

**Figure 4:**
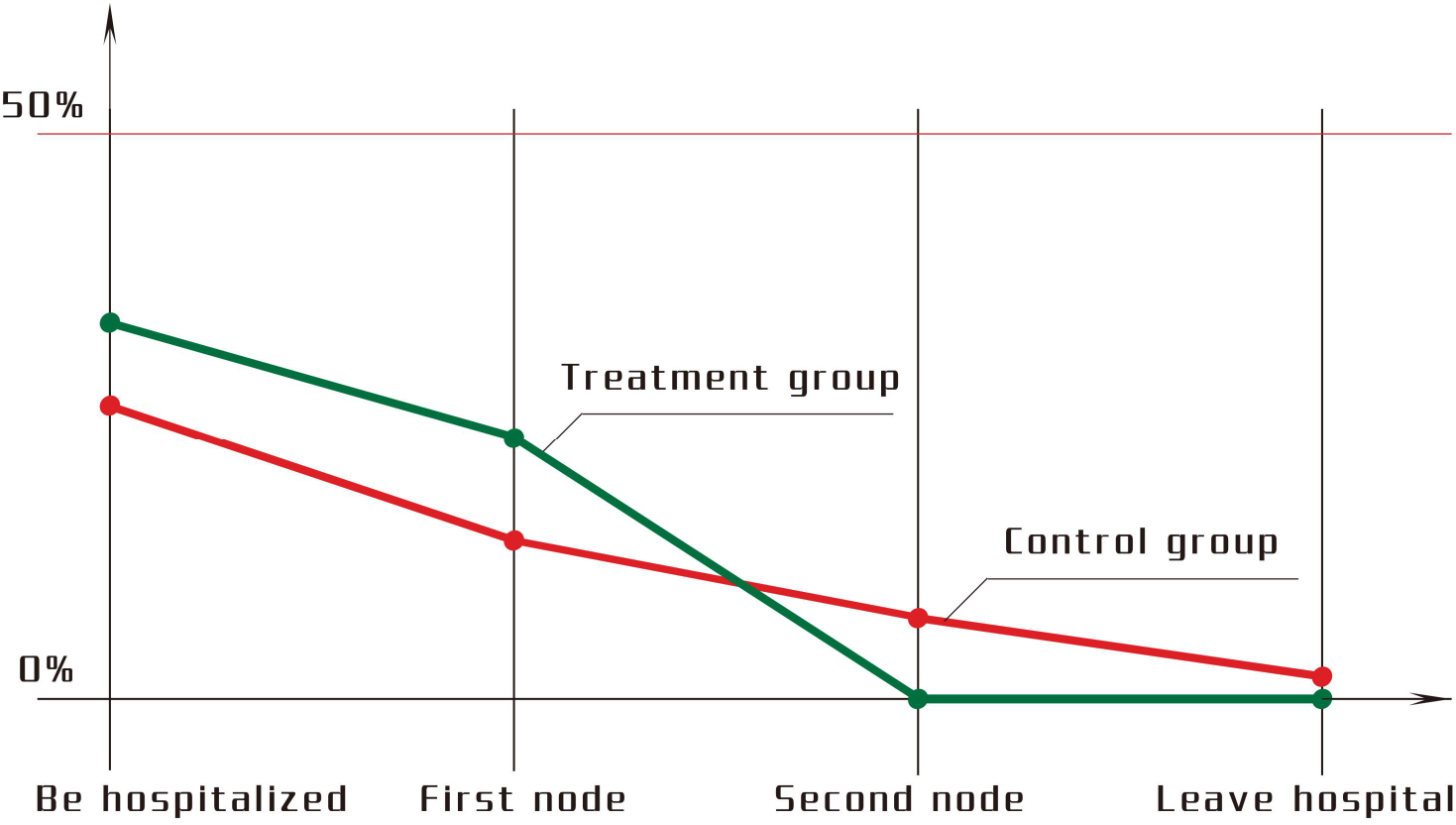
percentage change curve of patients of lymphocyte below normal in two groups

### Analysis of rehabilitation speed in two groups

A total of 20 patients took 3 bottles of high fiber whey short peptide as enteral nutrition preparation every day and were chosen as a treatment group of 3 bottles with one child excluded. The median date of treatment group of 3 bottles was March 4.35, 2020 as the reference point in time. After March 4, 2020, whole protein was used as enteral nutrition intervention every day till discharged from hospital. A total of 65 patients were used as a control group to compare rehabilitation speed. The average length of stay in the control group after March 4 was 6 days, minus the median standard time difference of 0.47 days, as the average speed of turning negative in the control group. There was no significant difference between general situation of the two groups (P > 0.05).The rehabilitation speed of the treatment group was significantly faster than that of the control group (P = 0.015) (Table 8).

**Table 8.** Comparison of the speed of turning Yin between two groups

| node | Treatment group(n=19) | control group(65) |
| --- | --- | --- |
| gender(man/woman) | 9/10 | 29/36 |
| average age/year | 49±18.68 | 56.2±14.6 |
| Pneumonia type (general / severe) | 16/3 | 42/23 |
| Maximum turn negative time | 13 | 14 |
| Use » 10 days number of people | 2 | 15 |
| Number of people in 1 day | 6 | 8 |
| Average transit parameters (days) | 3.37±3.22 | 6±4.25 |
| Average time of turning negative | 3.37 | 5.53 |
| Average time of turning negative | 60.94% |  |

One bottle of high fiber whey short peptide per day was taken as enteral nutrition intervention for all 12 common patients, as a 1-bottle treatment group.With the same comparison method,there was no significant difference between general situation of the two groups (P > 0.05),or between rehabilitation speed of the two groups (P = 0.836) (Table 9).

**Table 9.** Comparison of the speed of turning negative in one bottle groups

| node | Treatment | control group(37) |
| --- | --- | --- |
| gender(man/woman) | 7/5 | 15/22 |
| average age/year | 48±18.68 | 54.84±14.6 |
| Pneumonia type (general ) | 12 | 37 |
| Maximum turn negative time | 11 | 14 |
| Use » 10 days number of people | 3 | 11 |
| Number of people in 1 day | 1 | 4 |
| Average transit parameters (days) | 6.00±3.28 | 6.30±4.58 |
| Average time of turning negative (day) | 6 | 6.38 |
| Average time of turning negative | 94.04% |  |

Note: average speed of turning negative percentage: Mean time of turning negative in treatment.

## Discussion

SARS-CoV-2 infects the host using the angiotensin converting enzyme 2 receptor and causes covid-19, and this receptor is expressed in many organs, including the lungs and intestines.^1-5^Gene sequencing found that,the expression level of ACE2 receptor in digestive tract is almost 100 times of that in respiratory organs.^6^ACE2 and TMPRSS2 are the determinants of SARS-CoV-2 entry into cells, ACE2 is highly expressed in the absorptive intestinal epithelial cells, and the small intestine is rich furin protease. ^1-4^Electron microscopy revealed viral inclusion structures in endothelial cells of the blood vessels of patients with COVID-19, and in histological analyses, the small intestine showed endotheliitis (endothelialitis) of the submucosal vessels,and an accumulation of inflammatory cells associated with endothelium,as well as apoptotic bodies.^7^The vascular endothelium is an active paracrine,endocrine,and autocrine organ that is indispensable for the maintenance of vascular homoeostasis.^8^Endothelial dysfunction is a principal determinant of microvascular dysfunction by shifting the vascular equilibrium towards more vasoconstriction with subsequent organ ischaemia,inflammation with associated tissue oedema.^9^A large number of clinical studies have shown that 3% - 79% of patients with covid-19 have digestive system symptoms.^10-15^In this study,99.34% of patients with covid-19 were found to have prealbumin lower than norma at the time of admission,with an average of 149.28mg/L.Pathological,genetic and clinical data show that Sars-cov-2 uses ACE2 and TMPRSS2 to enter intestinal absorption cells and submucosal microvascular endothelial cells, causing diffuse inflammatory damage of gastrointestinal tract in patients with covid-19,which were mainly manifested in microvascular lesions under the intestinal mucosa and intestinal mucosal absorptive intestinal epithelial cell lesions,with serious impairment of digestive absorption function.^2-8^Short peptide predigested nutrition does not need to rely on human digestive function.In the treatment group, short peptide enteral nutrition intervention was used in the later stage of the course, and the prealbumin increased significantly in the treatment group. This finding is consistent with the results of pathology and gene examination, indicating that the patients with covid-19 have severe impairment of digestive absorption function.This finding provides a theoretical basis for the intervention of short peptide type predigested enteral nutrition in patients with covid-19.^16-20^ This finding is worth examining in the future research. The microvasculature under the small intestinal mucosa of patients with COVID-19 has serious inflammation and impaired digestion and absorption function,^5,6^which can not provide adequate nutrition for intestinal lymphocytes. Malnutrition can cause the decrease of systemic immune function.^34-45^More than 60% of patients with COVID-19 have lymphocytopenia,and almost all patients with COVID-19 have a decrease in the absolute number of T lymphocytes,CD4 + T cells and CD8 + T cells.^21-33^The negative nitrogen balance of patients with COVID-19 was 99.34%, and the proportion was consistent with the proportion of absolute number decrease of T lymphocytes,CD4 + T cells and CD8 + T cells in the literature. This suggests that there may be a direct relationship between nutritional status and immune function in patients with covid-19. In the later period of treatment the incidence of lymphocytopenia in the treatment group was significantly lower than that in the control group and literature reports.60% of human lymphocytes are distributed in the intestinal system.dipeptides and tripeptides can be actively absorbed, directly providing nutrition for intestinal lymphocytes,and improving the immune function of patients.^34-45^ These changes of prealbumin and lymphocytopenia in the treatment group may be the reason that the rehabilitation speed of the treatment group is significantly faster than that of the control group.

Most patients with COVID-19 developed severe anorexia at the time of admission.^9-15^The acceptance of patients is an important factor of oral nutritional supplements. Nano short peptide solves the taste problem of short peptide, and improves the acceptance of COVID-19 patients. It is an important factor for the successful implementation of this clinical nutrition treatment. In consideration of the limited number of cases in this study, we call for large-data double-blind controlled trials in the epidemic areas to examine these results carefully and provide a scientific basis for reducing the harm of the epidemic.

## Data Availability

All raw data are from Yichang Central People's Hospital, Hubei Province, China.

